# Impaired lung function and lung cancer risk in 461,183 healthy individuals: a cohort study

**DOI:** 10.1101/2023.10.29.23297726

**Authors:** Thu Win Kyaw, Min-Kuang Tsai, Chi-Pang Wen, Chin-Chung Shu, Ta-Chen Su, Xifeng Wu, Wayne Gao

## Abstract

**Background:** It has been known that smoking and various lung diseases including lung cancer can cause lung function impairment. However, the impact of different types of lung function impairments, such as preserved ratio impaired spirometry (PRISm) and airflow obstruction (AO), on the incidence and mortality of lung cancer in both general and never-smoker populations remains unclear. We wished to examine the effect of lung function impairments on lung cancer risks.

**Methods:** This was a retrospective cohort study of individuals from a health surveillance program in Taiwan who underwent baseline spirometry tests at the entry point. PRISm was defined as an FEV1/FVC (Forced Expiratory Volume in 1 second/ Forced Vital Capacity) ratio >0.7 and FEV1 <0.8, while AO was defined as an FEV1/FVC ratio <0.7. Cox proportional hazards models and cubic spline curves were used to examine the associations between lung function impairments and lung cancer risks.

**Results:** The study included 461,183 individuals, of whom 14.3% had PRISm and 7.9% had AO. A total of 4,038 cases of lung cancer and 3,314 lung cancer-related deaths were identified during the 23 years of follow-up. Individuals with PRISm and AO exhibited a higher risk of lung cancer incidence and mortality compared to those with normal lung function. The adjusted hazard ratios (aHRs) and 95% confidence intervals (95%CI) were 1.14 (1.03-1.26) and 1.23 (1.10-1.37) in the overall cohort, and 1.08 (0.93-1.24), and 1.23 (1.05-1.45) in the never-smoker cohort. The risks of both developing and dying of lung cancer increased with the severity levels of lung function impairments and lower FEV1 values.

**Conclusion:** Impaired lung function is associated with increased risks of developing lung cancer and subsequent mortality. The study highlights the importance of considering lung function in lung cancer screening for better candidate selection.

**WHAT IS ALREADY KNOWN ON THIS TOPIC:** Impaired lung function is a common condition that can be observed in people with smoking habits and other respiratory illnesses including lung cancer. However, the effect of lung function impairment alone on the risks of lung cancer incidence and mortality is not clear. Early detection of lung cancer is essential for effective disease management, and lung cancer screening is a key preventive measure that can help achieve this. However, current lung cancer screening guidelines only consider age and smoking history, not lung function status.

**WHY THIS STUDY ADDS:** Impaired lung function is associated with an increased risk of lung cancer in a large Asian cohort. Nonetheless, among never-smoking individuals exhibiting spirometrically defined PRISM/AO, the observed risks were found to lack statistical significance, with the exception of lung cancer mortality within the non-smoking PRISm subgroup.

**HOW THIS STUDY MIGHT AFFECT RESEARCH, PRACTICE OR POLICY:** Lung function status should be taken into consideration in lung cancer screening criteria. The importance of monitoring and addressing lung function impairment in lung cancer risk management should also be widely shared with the medical community and the public.

## INTRODUCTION

Lung cancer is the deadliest cancer, with the highest mortality rate and second-highest incidence rate. In 2020 alone, it accounted for 2,206,771 new cases (11.4%) and caused 1,796,144 deaths (18%) worldwide.[1] Cigarette smoking stands out as the most significant contributor, being responsible for approximately 80-90% of all lung cancer cases.[2] Other well-known risk factors include exposure to carcinogens like tobacco smoke, radon, and air pollution, as well as pre-existing pulmonary conditions such as chronic obstructive pulmonary disease (COPD) and age-related intrinsic factors.[3] Lung function status is not taken into consideration in current screening criteria which are based only on age and pack-year smoking history.[4]

It is well-recognized that individuals with smoking habits, lung cancer, or respiratory illnesses have impaired lung function. For example, TB and asthma are risk factors for lung function impairment among Korean non-smokers[5], and air pollution, even at very low levels, has adverse effects on lung function in adults.[6] There have been studies on lung function and lung cancer however most studies targeted smokers or patients with COPD or PRISm or lung cancer. A retrospective study of COPD infradiagnosis in lung cancer (RECOIL) study reported that COPD is often underdiagnosed in patients with lung cancer and this can lead to more advanced cancer stages and possibly worse outcomes.[7] The severity of COPD is independently associated with the risk of developing lung cancer[8] and smoking status modifies this association.[9] Another form of lung function impairment, the restrictive type PRISm, was also linked to an elevated mortality risk. This association appears unrelated to smoking, obesity, or pre-existing lung disease, despite PRISm being transient in many patients.[10] The prevalence of PRISm also was as high as 17-24% in a retrospective analysis of over twenty thousand spirometries.[11]

The FEV1 decline rate may be a biomarker for developing lung cancer.[12] It may still be a predictor of lung cancer, especially for women,[13] however, genetically decreased FEV1 is not causally correlated with lung cancer incidence.[13] In small cell lung cancer (SCLC) patients, a low FEV1, not COPD, is a predicting factor for poor treatment outcomes.[14] Some studies, however, have proposed that the variance in lung function measures among smokers is not fully explained by the smoking intensity and the number of pack-years smoked did not affect the lung cancer rate in patients with COPD.[15]

A study in 2017[16] said the annual frequency of lung cancer in never-smokers had more than doubled in the past 7 years, from 13% to 28%. This increase was due to an absolute increase in the number of never-smokers developing lung cancer, not simply a change in the ratio of never-smokers to current and former smokers. Patients with lung cancer who had never smoked typically presented with non-specific symptoms and most were detected on incidental imaging.[16] A previous study found that the proportion of non-smoking patients with non-small cell lung cancer (NSCLC) increased between 1990 and 2013. This increase was seen in a large, diverse patient population and independent of sex, stage, and race/ethnicity. This suggests that the actual incidence of lung cancer in never smokers is increasing.[17] A UK Biobank study on 222,274 never-smokers aged 40-69 years with 4 years of follow-up found that lung function can be a modest predictor of lung cancer risk in never-smokers, however, there acknowledged the potential selection bias of containing more male and white participants in the study cohort.[18] Overall, the relationship between lung function and lung cancer is complex and multifactorial.

We hypothesized that the presence of impaired lung function statuses alone may be an independent risk factor for lung cancer. The objective of our study was to investigate the potential causal relationship between lung function impairments, particularly restricted type (PRISm), and lung cancer risks. The implication of our study was to suggest the inclusion of lung function status in future lung cancer screening criteria.

## METHOD

### Study Design & Participants

It was a retrospective cohort utilizing baseline data of participants enrolled in a health screening program in Taiwan between 1994 and 2017. Participants were followed up for lung cancer development and death until December 2017.

This study included 646,987 healthy individuals (aged ≥ 20). Getting signed consent forms, baseline data on demographics, lifestyle, and medical history were collected using a self-administered questionnaire. A standardized physical examination and laboratory tests were performed including spirometry for assessing lung function using HI-501, HI-701, or HI-801 spirometers from Chest M.I. Inc, Tokyo, Japan. Results were measured as percentages of FEV1 and FVC. Participants were excluded based on criteria of missing values of FEV1 or FVC (n =109,122), FEV1/FVC >1 (n =71,818), FEV1 or FVC percentage >150 or <25 (n =4,864), and already diagnosed with lung cancer (n =152). The final cohort consisted of 461,031 participants. (Figure 1) Ethical approval for this study was obtained from Institutional Review Board at the China Medical University (Approval Number: CMUH111-REC2-118). All participants gave consent to analyze their unidentifiable data before being included in this study. All data analyses were conducted at the Data Science Center of the Ministry of Health and Welfare in Taiwan, where all data were de-identified to protect participant confidentiality.

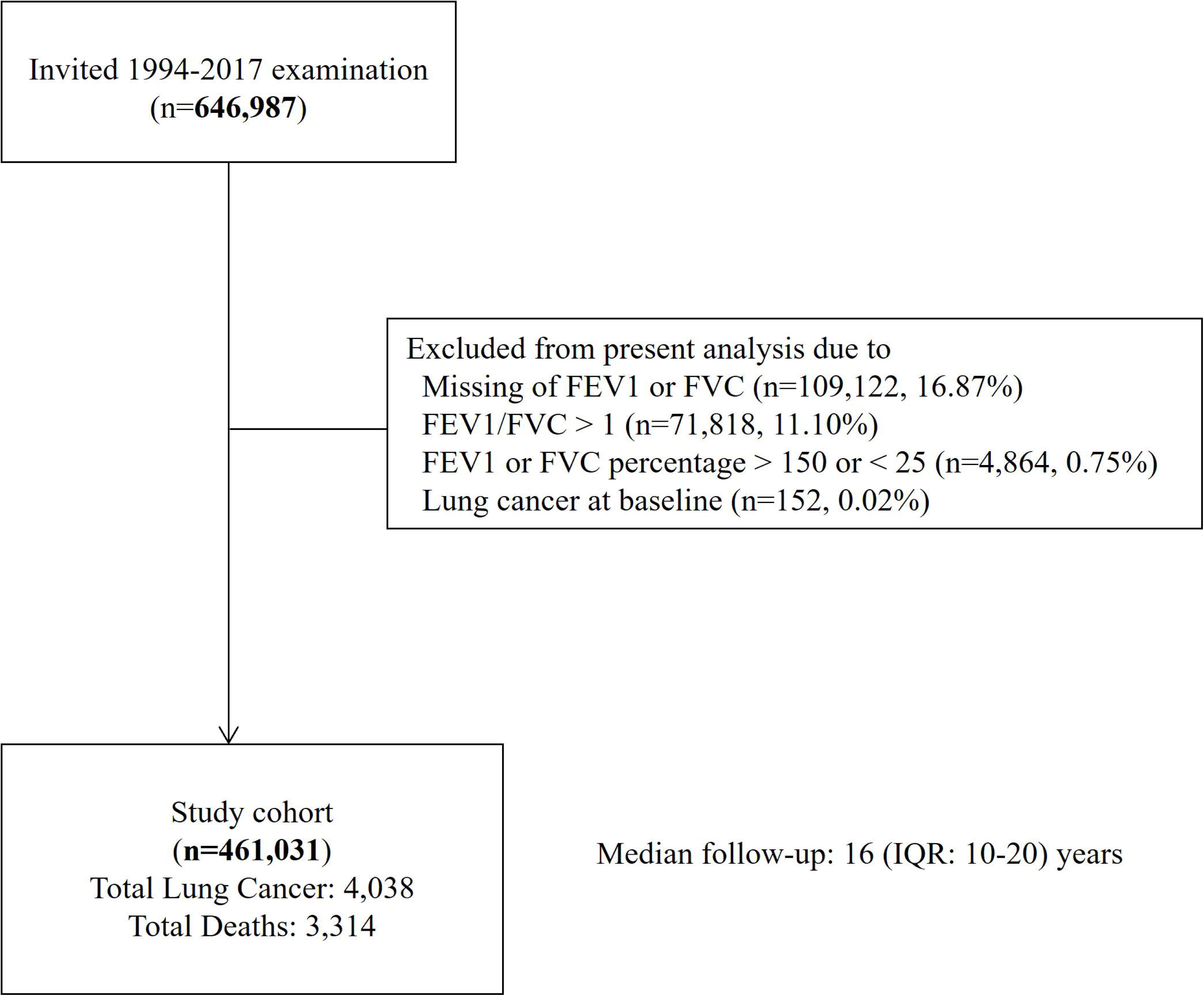
Figure 1.

### Variables and definitions

Based on spirometry results, participants were classified into normal, PRISm, and AO. PRISm was defined as FEV1/FVC ratio >0.7 and FEV1 <80% predicted, AO as FEV1/FVC ratio <0.7, and normal as FEV1/FVC ratio >0.7 and FEV1 >80% predicted. Severity levels of PRISm[19] were determined based on FEV1 values: >70, 60-69, 50-59, and <50. For AO, Global Initiative for Chronic Obstructive Lung Disease (GOLD) stages I-IV were applied, stage I (mild) for FEV1 values >80, II (moderate) for 50-79, III (severe) for 30-49, and IV (very severe) for <30.

The primary outcomes of the study were newly diagnosed lung cancer cases after the entry time point and lung cancer-related deaths during the follow-up period by matching patients’ identification numbers with the Nationwide Registry of Patients with Catastrophic Illness and National Death File until December 2017. Causes of death were coded according to the International Classification of Diseases, 9th version (ICD-9).

Several covariates were adjusted during analyses. Diabetes and hypertension cases were identified based on medical history or positive screening results such as fasting blood glucose 126 mg/dL or higher and systolic blood pressure 140 mmHg or higher. Other variables included age, sex, educational levels, smoking status, drinking status, body mass index (BMI), and physical activity.

### Statistical processing and analysis

Descriptive statistics were used to tabulate the baseline characteristics of participants by lung function impairment categories. The Multivariable Cox proportional hazards model was applied to investigate the relationship between lung function impairment and the risks of lung cancer incidence and mortality. Hazard ratios (HRs) and 95% confidence intervals (CIs) were calculated, adjusting for confounding variables of age, sex, education level, smoking status, alcohol consumption, BMI, physical activity level, hypertension, and diabetes status.

To ensure the credibility of our findings and to eliminate any potential bias arising from reverse causation, we conducted sensitivity analyses. We analyzed specific subgroups by excluding individuals who smoked and those who developed lung cancer within 3 years of the health screening program entry. For instance, the exclusion of participants with baseline hypertension in a study from Hong Kong[20] and the exclusion of participants with asthma in the UK Biobank study[10]. Stratified analyses by age, gender, smoking status, and histological cell types were also performed. Furthermore, hazard ratios were plotted against continuous values of FEV1 on cubic spline curves to observe the correlation trends. SAS analytical software version 9.4 was applied, and all tests were two-sided with a significance level of α=0.05.

### Patient and public involvement

This analysis was based on anonymized retrospective data from a health surveillance program in Taiwan.

## RESULTS

Out of 461,031 individuals, 14.3% had PRISm, 7.9% had AO, and 77.8% had normal lung function. Among the never-smoker subgroup, 13.8% had PRISm, 3.5% had AO, and 82.7% had normal lung function. During the 23-year follow-up period of the study, 4,038 cases of lung cancer and 3,314 lung cancer-related deaths were identified. The incidence rate of lung cancer was 28.55 cases per 100,000 person-years, and the mortality rate was 21.90 cases per 100,000 person-years.

### Characteristics of Participants by lung function statuses

The main characteristics of the study participants by lung function statuses are shown in Table 1. In comparison to the AO group, the individuals in the PRISm group were relatively older age i.e., over 40 and more likely to be females, never-smokers, non-drinkers, higher BMI i.e., above 25, physically less active, with comorbidities of diabetes and hypertension. However, there were no significant differences in respiratory symptoms between the two groups. Characteristics of the study subjects by severity levels of restrictive lung condition (PRISm) were presented in Supplementary Table 1.

**Table 1.** Characteristics of participants by lung function status.

|  |  | Total | Normal lung function | PRISm | COPD |
| --- | --- | --- | --- | --- | --- |
|  |  | N* (%) | N* (%) | N* (%) | N* (%) |
|  |  | 461,031 (100.0) | 358,755 (77.8) | 65,781 (14.3) | 36,495 (7.9) |
| Age | 20-39 | 239,283 (51.9) | 207,047 (57.7) | 17,563 (26.7) | 14,673 (40.2) |
|  | 40-59 | 158,298 (34.3) | 117,975 (32.9) | 28,406 (43.2) | 11,917 (32.7) |
|  | 60 or above | 63,450 (13.8) | 33,733 (9.4) | 19,812 (30.1) | 9,905 (27.1) |
| Gender | Men | 225,587 (48.9) | 178,160 (49.7) | 28,853 (43.9) | 18,574 (50.9) |
|  | Women | 235,444 (51.1) | 180,595 (50.3) | 36,928 (56.1) | 17,921 (49.1) |
| Education | Middle school or below | 88,366 (25.0) | 57,273 (19.7) | 24,577 (50.9) | 6,516 (45.7) |
|  | High school | 81,725 (23.1) | 68,519 (23.6) | 10,055 (20.8) | 3,151 (22.1) |
|  | Junior college | 69,955 (19.8) | 62,019 (21.3) | 6,004 (12.4) | 1,932 (13.5) |
|  | College or above | 113,121 (32.0) | 102,773 (35.4) | 7,688 (15.9) | 2,660 (18.7) |
| Smoking status | Never-smoker | 241,874 (69.9) | 199,917 (70.0) | 33,466 (71.2) | 8,491 (63.2) |
|  | Ex-smoker | 22,879 (6.6) | 18,524 (6.5) | 3,148 (6.7) | 1,207 (9.0) |
|  | Current smoker | 81,120 (23.5) | 66,961 (23.5) | 10,418 (22.2) | 3,741 (27.8) |
| Drinking status | Non-drinker | 266,918 (78.7) | 221,936 (79.2) | 35,475 (77.7) | 9,507 (71.4) |
|  | Occasional drinker | 39,478 (11.6) | 32,820 (11.7) | 4,689 (10.3) | 1,969 (14.8) |
|  | Regular drinker | 32,641 (9.6) | 25,300 (9.0) | 5,510 (12.1) | 1,831 (13.8) |
| Body mass index | <18.5 | 33,308 (7.2) | 26,577 (7.4) | 4,406 (6.7) | 2,325 (6.4) |
|  | 18.5~24 | 293,049 (63.6) | 234,579 (65.4) | 35,026 (53.3) | 23,444 (64.3) |
|  | 25~29 | 114,257 (24.8) | 83,464 (23.3) | 21,552 (32.8) | 9,241 (25.3) |
|  | ≥30 | 20,299 (4.4) | 14,086 (3.9) | 4,739 (7.2) | 1,474 (4.0) |
| Physical activity | Inactive | 181,794 (51.1) | 148,488 (50.9) | 26,108 (53.1) | 7,198 (49.9) |
|  | Low | 85,563 (24.1) | 72,244 (24.7) | 10,051 (20.4) | 3,268 (22.7) |
|  | Moderate | 53,494 (15.0) | 43,729 (15.0) | 7,664 (15.6) | 2,101 (14.6) |
|  | High | 21,816 (6.1) | 17,013 (5.8) | 3,565 (7.2) | 1,238 (8.6) |
|  | Very high | 12,955 (3.6) | 10,514 (3.6) | 1,819 (3.7) | 622 (4.3) |
| Hypertension | None | 367,272 (79.7) | 298,479 (83.2) | 42,021 (63.9) | 26,772 (73.4) |
|  | Yes | 93,759 (20.3) | 60,276 (16.8) | 23,760 (36.1) | 9,723 (26.6) |
| Diabetes | None | 435,832 (94.5) | 343,469 (95.7) | 58,235 (88.5) | 34,128 (93.5) |
|  | Yes | 25,199 (5.5) | 15,286 (4.3) | 7,546 (11.5) | 2,367 (6.5) |

\* N Number of participants
PRISm: Preserved Ratio Impaired Spirometry; COPD: Chronic Obstructive Pulmonary Disease.

### Risks of lung cancer and lung function impairments

Findings for the overall cohort in Table 2 showed individuals diagnosed with PRISm exhibited a 14% higher risk of developing lung cancer and a 23% higher risk of dying from lung cancer compared to individuals with normal lung function. Similarly, individuals with AO had a 29% increased risk of lung cancer incidence and a 30% higher risk of lung cancer mortality. Notably, the risks of developing lung cancer were found to escalate in proportion to the severity levels of lung function impairment.

**Table 2.**
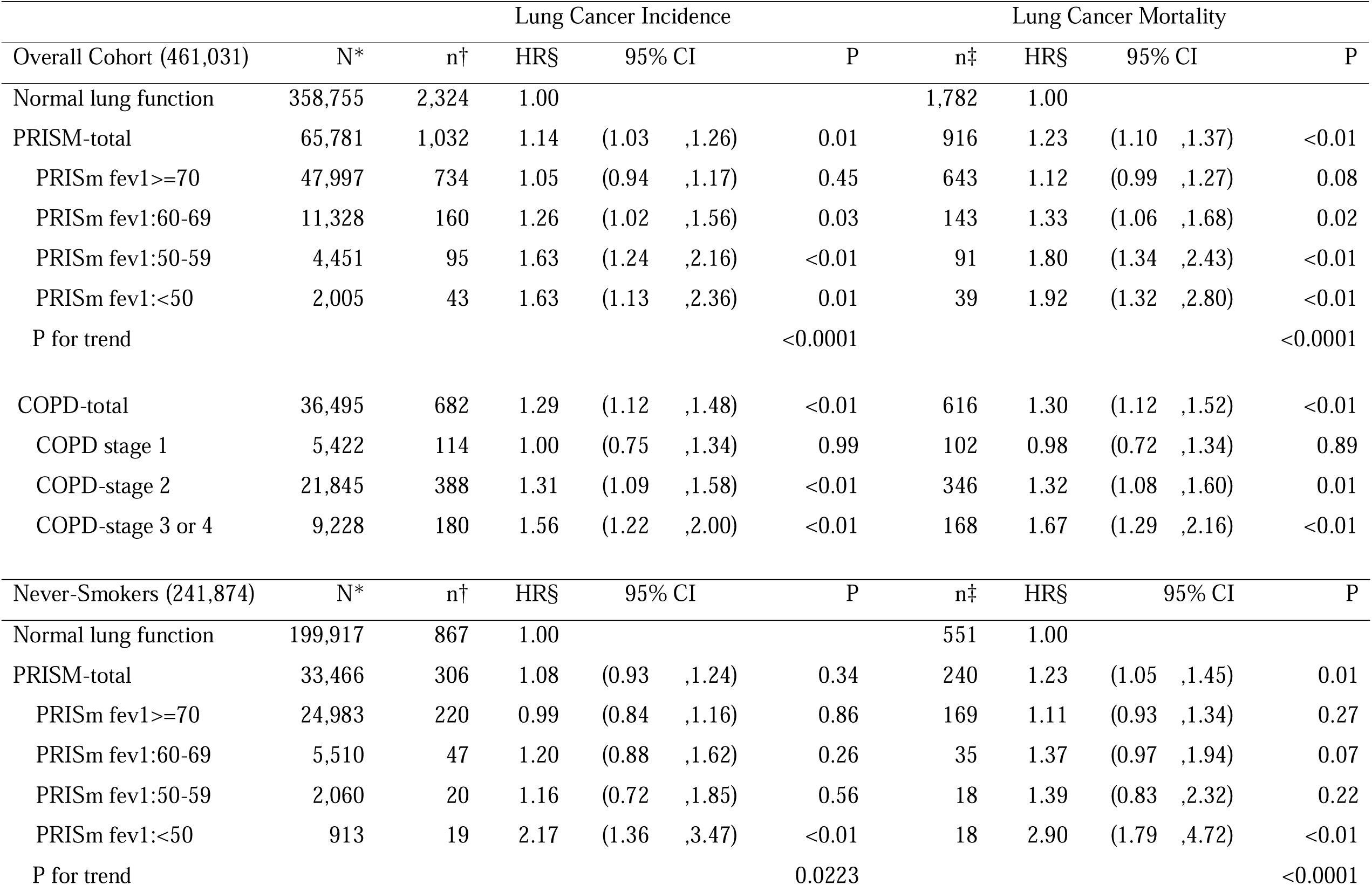
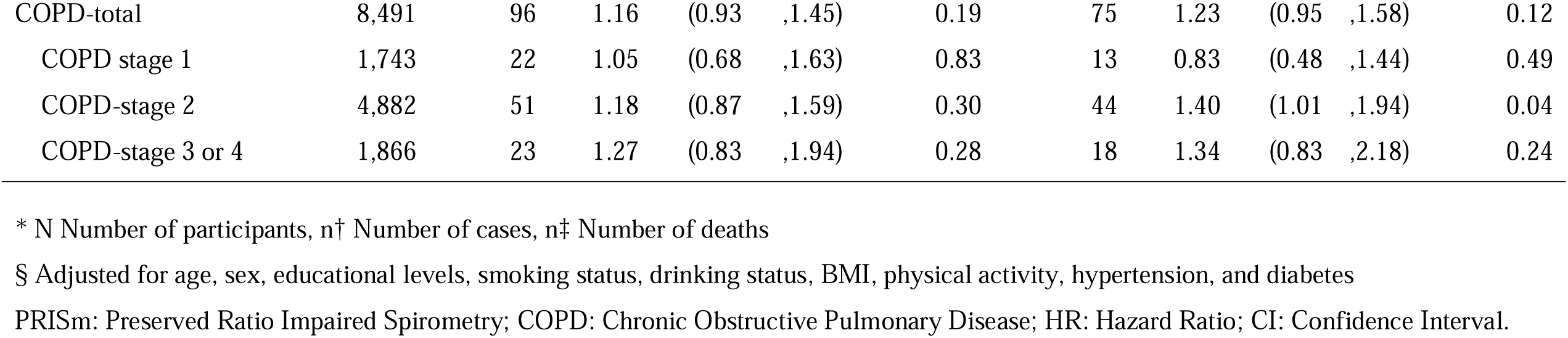
Risk of lung cancer incidence and mortality by lung function impairment severities in overall and never-smokers cohorts.

| Lung Cancer Incidence |  |  |  |  |  |  | Lung Cancer Mortality |  |  |  |  |
| --- | --- | --- | --- | --- | --- | --- | --- | --- | --- | --- | --- |
| Overall Cohort (461,031) | N* | n† | HR§ | 95% CI |  | P | n‡ | HR§ | 95% CI |  | P |
| Normal lung function | 358,755 | 2,324 | 1.00 |  |  |  | 1,782 | 1.00 |  |  |  |
| PRISM-total | 65,781 | 1,032 | 1.14 | (1.03 | ,1.26) | 0.01 | 916 | 1.23 | (1.10 | ,1.37) | <0.01 |
| PRISm fev1>=70 | 47,997 | 734 | 1.05 | (0.94 | ,1.17) | 0.45 | 643 | 1.12 | (0.99 | ,1.27) | 0.08 |
| PRISm fev1:60-69 | 11,328 | 160 | 1.26 | (1.02 | ,1.56) | 0.03 | 143 | 1.33 | (1.06 | ,1.68) | 0.02 |
| PRISm fev1:50-59 | 4,451 | 95 | 1.63 | (1.24 | ,2.16) | <0.01 | 91 | 1.80 | (1.34 | ,2.43) | <0.01 |
| PRISm fev1:<50 | 2,005 | 43 | 1.63 | (1.13 | ,2.36) | 0.01 | 39 | 1.92 | (1.32 | ,2.80) | <0.01 |
| P for trend |  |  |  |  |  | <0.0001 |  |  |  |  | <0.0001 |
| COPD-total | 36,495 | 682 | 1.29 | (1.12 | ,1.48) | <0.01 | 616 | 1.30 | (1.12 | ,1.52) | <0.01 |
| COPD stage 1 | 5,422 | 114 | 1.00 | (0.75 | ,1.34) | 0.99 | 102 | 0.98 | (0.72 | ,1.34) | 0.89 |
| COPD-stage 2 | 21,845 | 388 | 1.31 | (1.09 | ,1.58) | <0.01 | 346 | 1.32 | (1.08 | ,1.60) | 0.01 |
| COPD-stage 3 or 4 | 9,228 | 180 | 1.56 | (1.22 | ,2.00) | <0.01 | 168 | 1.67 | (1.29 | ,2.16) | <0.01 |
| Never-Smokers (241,874) | N* | n† | HR§ | 95% CI |  | P | n‡ | HR§ | 95% CI |  | P |
| Normal lung function | 199,917 | 867 | 1.00 |  |  |  | 551 | 1.00 |  |  |  |
| PRISM-total | 33,466 | 306 | 1.08 | (0.93 | ,1.24) | 0.34 | 240 | 1.23 | (1.05 | ,1.45) | 0.01 |
| PRISm fev1>=70 | 24,983 | 220 | 0.99 | (0.84 | ,1.16) | 0.86 | 169 | 1.11 | (0.93 | ,1.34) | 0.27 |
| PRISm fev1:60-69 | 5,510 | 47 | 1.20 | (0.88 | ,1.62) | 0.26 | 35 | 1.37 | (0.97 | ,1.94) | 0.07 |
| PRISm fev1:50-59 | 2,060 | 20 | 1.16 | (0.72 | ,1.85) | 0.56 | 18 | 1.39 | (0.83 | ,2.32) | 0.22 |
| PRISm fev1:<50 | 913 | 19 | 2.17 | (1.36 | ,3.47) | <0.01 | 18 | 2.90 | (1.79 | ,4.72) | <0.01 |
| P for trend |  |  |  |  |  | 0.0223 |  |  |  |  | <0.0001 |

|  |  |  |  |  |  |  |  |  |  |
| --- | --- | --- | --- | --- | --- | --- | --- | --- | --- |
| COPD-total | 8,491 | 96 | 1.16 | (0.93 ,1.45) | 0.19 | 75 | 1.23 | (0.95 ,1.58) | 0.12 |
| COPD stage 1 | 1,743 | 22 | 1.05 | (0.68 ,1.63) | 0.83 | 13 | 0.83 | (0.48 ,1.44) | 0.49 |
| COPD-stage 2 | 4,882 | 51 | 1.18 | (0.87 ,1.59) | 0.30 | 44 | 1.40 | (1.01 ,1.94) | 0.04 |
| COPD-stage 3 or 4 | 1,866 | 23 | 1.27 | (0.83 ,1.94) | 0.28 | 18 | 1.34 | (0.83 ,2.18) | 0.24 |

\* N Number of participants, n† Number of cases, n‡ Number of deaths
§ Adjusted for age, sex, educational levels, smoking status, drinking status, BMI, physical activity, hypertension, and diabetes
PRISm: Preserved Ratio Impaired Spirometry; COPD: Chronic Obstructive Pulmonary Disease; HR: Hazard Ratio; CI: Confidence Interval.

### Lung cancer risks by lung function impairments in sensitivity analyses

After excluding smoking individuals, the risks of developing lung cancer were still higher among those with impaired lung function. Analyses for the subgroup of never-smokers in Table 2 revealed individuals with PRISm had an 8% higher chance of developing lung cancer and a 23% higher chance of dying from lung cancer compared to those with normal lung function. Similarly, for individuals with AO, the increased risks were 16% for lung cancer incidence and 23% for lung cancer mortality. These findings remained consistent even when lung cancer cases occurring within the first 3 years were excluded to avoid reverse causation bias, as demonstrated in Supplementary Figure 1 (a) and (b).

### Lung cancer risks by lung function impairments in subgroups analyses

Stratification on the individuals with PRISm by age, gender, and smoking status, individuals with impaired lung function were consistently found to have elevated risks of both lung cancer incidence and mortality. (Figure 2) Those aged over 60 had higher risks compared to those under 60. Females exhibited a higher risk of lung cancer mortality compared to males. Current and ex-smokers had a higher risk of lung cancer incidence compared to never-smokers. In stratified analyses by histological types of lung cancer for never-smokers, the risks of developing or dying from squamous cell carcinoma were found to be higher than those of adenocarcinoma. (Supplementary Figure 2)

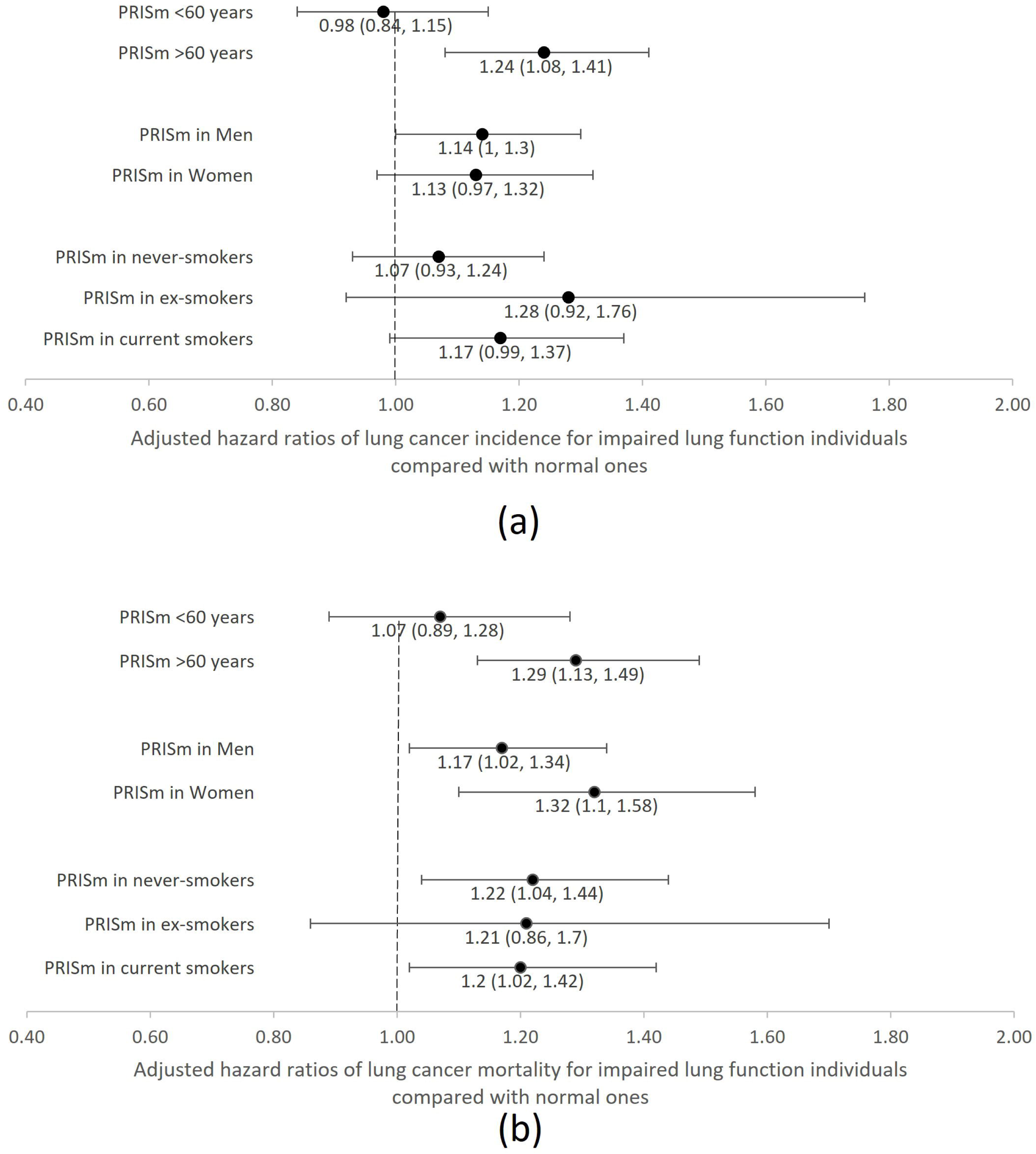
Figure 2.

### Relationship between continuous values of FEV1 and hazard ratios

Cubic spline models were employed to analyze the correlation between FEV1 values and the likelihood of lung cancer incidence and mortality within the PRISm group of never-smokers. The findings from Figure 3 revealed a noteworthy inverse relationship, suggesting that under the predicted FEV1 80%, individuals with lower FEV1 values had elevated risks of developing and dying from lung cancer. Under the FEV1 value of 50%, never-smokers with PRISm exhibited significantly higher risks for both lung cancer incidence and mortality. Moreover, the slopes of cubic spline curves in squamous cell carcinoma were steeper, indicating greater risks, compared to adenocarcinoma. (Figures not shown)

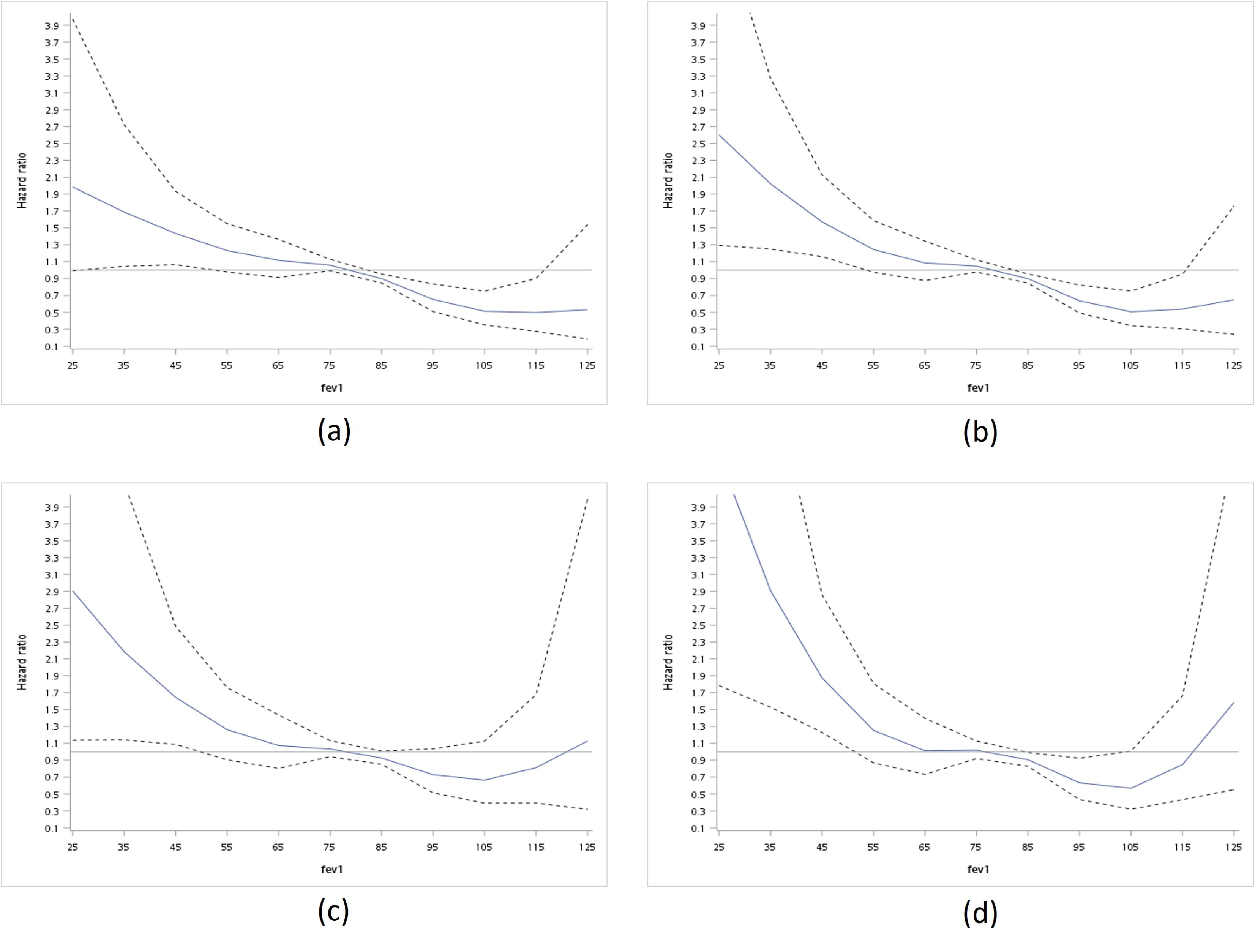
Figure 3.

## DISCUSSION

Our study found that people with lung function impairments, such as PRISm and AO, have higher risks of lung cancer incidence and mortality. The risks of lung cancer increased as the severity of lung function impairment increased or as the FEV1 values lowered. These findings of dose responsiveness and reverse correlation remained consistent across sensitivity analyses, stratified analyses, and cubic spline curves, demonstrating the robustness of the association.

In our study, individuals with AO had a higher risk of lung cancer incidence and mortality than individuals with PRISm. This could be attributed to the larger number of cases in low severity levels (FEV1 >70%) of PRISm-type lung function impairment. At high severity levels (FEV1 value <50%), individuals with PRISm faced higher risks compared to those with AO. Our findings are consistent with the report published by Young et al. in 2022, which found that individuals with PRISm have more advanced lung cancers and the greatest lung cancer lethality.[21]

The prevalence of PRISm and AO in previous population-based and hospital-based studies gave a wide range of results. In our study, the proportions of PRISm and AO were 14.3% and 7.9%, compared to 10.2% and 13.1% in a population-based cohort in Korea.[22] These differences could be attributed to variations in the study population, such as a higher number of low-risk which means never-smokers in our cohort. Higher proportions of PRISm could be found in hospital-based or cohorts of high-risk groups. It was as high as 21.3% in the Korean lung cancer registry[23], 16.7% in a Japanese study of individuals over 40 years of age[24], and 17-24% in a hospital study from the University of Iowa[11]. Restrictive patterns of lung function impairment can be observed in interstitial lung diseases such as idiopathic pulmonary fibrosis (IPF). However, the prevalence of IPF is relatively low in Taiwan i.e., 3.1 to 6.4 cases per 100,000 people[25]. Therefore, the restrictive patterns observed in our cohort were more likely due to PRISm or AO rather than underlying interstitial lung diseases. Additionally, our study’s cohort was based on an apparently healthy general population enrolled in a private health screening program and we excluded extreme values of spirometry test readings (FEV1 or FVC percentage > 150 or < 25) for analyses.

In our study, the prevalence of AO was 7.9%, which is close to 8.8% reported for the South-East Asia Region/Western Pacific Region in a meta-analysis.[26] However, a higher prevalence of AO 37.4% was observed in a Manchester study targeted on ever-smokers aged 55-74.[27] Approximately one-third of smokers eligible for lung cancer screening in Germany had AO or PRISm[28], and 34.4% of the National Lung Screening Trial-American College of Radiology Imaging Network (NLST-ACRIN) cohort had COPD[29]. Importantly, the prevalence of AO can vary depending on the cutoff value used for diagnosis. Using a pre-bronchodilator FEV1/FVC<0.66 instead of 0.7 resulted in a 15% increase in the accuracy of AO diagnosis.[30] Despite these variations, our study’s findings agree with previous literature on the increasing lung function impairments in the general population, either PRISm or AO type.

The risks of lung cancer incidence and mortality for individuals with PRISm or AO in previous literature were higher than our study results. However, it is important to note that most of those studies were not based on general populations. In the overall cohort of our study, individuals with PRISm had a 14% higher chance of developing lung cancer which is relatively lower compared to the 27% higher risk observed in a nationwide population-based cohort study conducted in Korea[22] Additionally, a Swedish study on construction workers reported a relative risk (RR) of 2.0 (95%CI 1.6 – 2.5).[31] Individuals with AO in our study had a 29% higher risk for lung cancer, however, the aforementioned Korean study reported two and half times higher risk in their analysis.[22] In a Norway study conducted on former or current smokers between the ages of 40 and 76 years, the risk of developing lung cancer for AO patients was reported to be as high as five times.[32]

Individuals with PRISm in our overall cohort had a 23% higher risk for lung cancer mortality. In the population-based Tucson Epidemiological Study of Airway Obstructive Disease (TESAOD) study of the 2048 cohort, participants with recurrent PRISm had 30% lower risk and inconsistent PRISm had 50% higher risk but the hazard ratios were not significant due to a low number of cases identified.[33] Non-small cell and small cell lung cancer patients with PRISm had poorer overall survival with a 62% higher risk of mortality, compared to those with AO or normal lung function.[23] In our cohort study, individuals with AO had a 30% higher risk of lung cancer mortality which was relatively low compared to other studies. For instance, in studies conducted on lung cancer patients, AO was associated with poorer overall and progression-free survivals with more than two times higher mortality risks.[34] Our study generally agreed with findings from previous literature and relatively weaker strength of associations could be explained by differences in cohort sizes, study design, and analytical settings.

In sensitivity analyses that excluded smokers and lung cancer cases occurring within the initial three-year period, we observed associations between the risk of lung cancer incidence and impairments in lung function. However, the risks were not significant among never-smokers with PRISm/AO, except for lung cancer mortality within the PRISm subgroup. The relatively small numbers of lung cancer cases and deaths among never-smokers in the cohort may explain the lack of significance observed in the analysis. We believe this finding is still relevant because even though no proven direct association, the UK biobank study[18] acknowledged the advantage of identifying lung cancer cases in never-smokers by including lung function status. The cubic spline curves in our study also showed the risk of lung cancer increased with the severity of lung function impairment in never-smokers with PRISm. These dose-response shape and inverse correlation were more obvious in people with severely reduced FEV1 values i.e., <50%. (Figure 3)

Our stratified analyses revealed that PRISm at age <60 had lower risks for both lung cancer incidence and mortality compared to those ages>60. However, the result was insignificant with 95% confidence intervals of overriding 1. This may be attributed to the low number of lung cancer cases and fatalities in the age group under 60. The Okinawa COPD casE finding AssessmeNt (OCEAN) study in Japan showed that a significant proportion of individuals aged <60 who attended their annual health examination had impaired lung function.[24] Similarly in our study, 70% of those with PRISm or AO were under 60 and were potentially engaging with respiratory illnesses including lung cancer in the future. In terms of stratification by gender and lung cancer histological types, it was observed that women exhibited a greater mortality risk from lung cancer in comparison to men. Furthermore, non-smoking individuals with PRISm demonstrated higher risks of squamous cell carcinoma than adenocarcinoma. However, the German Lung Cancer Screening Intervention Study (LUSI) shed light on the reasons behind the greater reduction in lung cancer mortality through CT screening in women, explaining the higher prevalence of slow-progressing and peripherally located adenocarcinomas in females, whereas males exhibited a greater association with rapidly growing and centrally located squamous cell carcinomas.[28]

The inverse relationship between FEV1 values (<80%) and lung cancer risks observed under cubic spline curves for nonsmoking individuals with PRISm is also consistent with previous literature concluding the association of rapid FEV1 decline with lung cancer development.[12, 13] FEV1 <80% is associated with inferior clinical outcomes and is also an independent risk factor for shorter overall survival in non-small cell lung cancer (NSCLC) patients.[35, 36] Furthermore, the outcomes of lung cancer patients with ongoing treatment can be estimated through lung function status.[34, 37]

The lungs’ primary function is to exchange gases, and the quality of the air or substance being breathed in is very important for health. Inhaled hazardous substances can damage cilia which then fail to propel trapped pathogens,[38] leading to prolonged exposure and eventually reducing respiratory function. In the absence of external factors, impaired lung function alone can initiate that vicious circle and consequently lung cancer-pro0moting events of inflammation, oxidative stress, DNA damage, fibrosis, and connective tissue deposition in the airways.[39] Genetic studies had also shown that pulmonary impairment can lead to lung cancer. Integrative analyses uncovered that pulmonary function can influence gene expression in lung tissue, which can then activate immune-related pathways that promote cancer growth.[40] The relationship between impaired lung function and lung cancer is complex, as both conditions are often caused by cigarette smoking. However, a recent study using Mendelian randomization analysis found that there is an independent link between lung function impairment and lung cancer risk, even after accounting for smoking. This suggests that there may be other factors, such as immune-related pathways, that contribute to the increased risk of lung cancer in people with impaired lung function.[41] According to our study results, the feasibility of including impaired lung function as a criterion for determining eligibility for lung cancer screening remains uncertain. Lung cancer screening did not have a mortality reduction benefit in people with severe airflow limitation.[21, 42] By incorporating FEV1 into screening criteria, we may preferentially select people who are not healthy enough to undergo follow-up curative treatments for lung cancer, or whose life expectancy is limited that they do not benefit from early detection of their cancer.

Strengths: Our study has several notable strengths. Firstly, it is a large cohort consisting of nearly half a million participants. Secondly, the long follow-up time of 23 years increased the statistical power and generalizability of findings. Thirdly, the study used a rigorous method to exclude those with pre-existing lung cancer before data analysis, reducing reverse causality bias. Fourthly, the robustness of the association was confirmed by consistent findings of association in sensitivity and stratified analyses. Finally, it is one of the rare studies that have examined the relationship between PRISm and AO for lung cancer incidence and mortality in the same cohort, with a particular focus on never-smokers.

Limitations: There exist also some limitations in our study. The study did not consider the possibility of transition to another type of lung function impairment, which may affect the findings. The study cohort was based on a self-paying private health surveillance program, mainly involving higher socioeconomic classes. The study cohort mainly represented residents in Taiwan, thus, the results may not be generalizable to other ethnicities or regions.

In conclusion, there is a significant association between impaired lung function and increased risks of lung cancer incidence and mortality, in the overall cohort. That relationship is independent of other potential confounding factors. However, the association was insignificant in the sub-cohort of never-smokers.

## Data Availability

All data produced in the present study are available upon reasonable request to the authors.

## Acknowledgment

The data used in this study was authorized by MJ Health Research Foundation (Authorization Code: MJHRFB2014001C). The MJ Health Research Foundation administered MJ Health Survey Database and MJ BioData, and the data were available at the website: http://www.mjhrf.org. The study design, data collection, data analysis, data interpretation, writing of the report, and submission for publication were independently decided by the authors and had no relation to the funding source. We are grateful to the Health and Welfare Data Science Center and National Health Research Institutes for providing administrative and technical support.

## Contributors

TWK, MKT, CPW, and WG conceptualized and designed the study. TWK and MKT analyzed and interpreted the data. TWK, MKT, and WG drafted and submitted the article for publication. TWK, MKT, CCS, TCS, CPW, XW, and WG critically revised the article for important intellectual content. All authors had final approval of the article.

## Patient and public involvement

Patients and/or the public were not involved in the design, conduct, reporting, or disseminating this research.

