## Supplementary Table 1 for "Impaired lung function and lung cancer risk in 461,183 healthy individuals: a cohort study"

Supplementary Table 1. Characteristics of participants by lung function status detailed in severity levels of PRISm

|  |  | Total | | Normal lung function | | Restricted lung | | Mild | | Moderate | | Moderately severe | | Severe or very severe | | AO | |
| --- | --- | --- | --- | --- | --- | --- | --- | --- | --- | --- | --- | --- | --- | --- | --- | --- | --- |
|  |  | N | (%) | N | (%) | N | (%) | N | (%) | N | (%) | N | (%) | N | (%) | N | (%) |
|  |  | 461,031 | (100.0) | 358,755 | (77.8) | 65,781 | (14.3) | 47,997 | (10.4) | 11,328 | (2.5) | 4,451 | (1.0) | 2,005 | (0.4) | 36,495 | (7.9) |
| Age | 20-39 | 239,283 | (51.9) | 207,047 | (57.7) | 17,563 | (26.7) | 11,937 | (24.9) | 3,795 | (33.5) | 1,358 | (30.5) | 473 | (23.6) | 14,673 | (40.2) |
|  | 40-59 | 158,298 | (34.3) | 117,975 | (32.9) | 28,406 | (43.2) | 20,974 | (43.7) | 4,825 | (42.6) | 1,829 | (41.1) | 778 | (38.8) | 11,917 | (32.7) |
|  | 60 or above | 63,450 | (13.8) | 33,733 | (9.4) | 19,812 | (30.1) | 15,086 | (31.4) | 2,708 | (23.9) | 1,264 | (28.4) | 754 | (37.6) | 9,905 | (27.1) |
| Gender | Men | 225,587 | (48.9) | 178,160 | (49.7) | 28,853 | (43.9) | 21,992 | (45.8) | 4,279 | (37.8) | 1,697 | (38.1) | 885 | (44.1) | 18,574 | (50.9) |
|  | Women | 235,444 | (51.1) | 180,595 | (50.3) | 36,928 | (56.1) | 26,005 | (54.2) | 7,049 | (62.2) | 2,754 | (61.9) | 1,120 | (55.9) | 17,921 | (49.1) |
| Education | Middle school or below | 88,366 | (25.0) | 57,273 | (19.7) | 24,577 | (50.9) | 18,337 | (50.7) | 3,751 | (47.8) | 1,628 | (55.1) | 861 | (62.0) | 6,516 | (45.7) |
|  | High school | 81,725 | (23.1) | 68,519 | (23.6) | 10,055 | (20.8) | 7,420 | (20.5) | 1,780 | (22.7) | 617 | (20.9) | 238 | (17.1) | 3,151 | (22.1) |
|  | Junior college | 69,955 | (19.8) | 62,019 | (21.3) | 6,004 | (12.4) | 4,435 | (12.3) | 1,074 | (13.7) | 352 | (11.9) | 143 | (10.3) | 1,932 | (13.5) |
|  | College or above | 113,121 | (32.0) | 102,773 | (35.4) | 7,688 | (15.9) | 5,946 | (16.5) | 1,236 | (15.8) | 359 | (12.1) | 147 | (10.6) | 2,660 | (18.7) |
| Smoking status | Never-smoker | 241,874 | (69.9) | 199,917 | (70.0) | 33,466 | (71.2) | 24,983 | (71.0) | 5,510 | (72.4) | 2,060 | (71.5) | 913 | (67.7) | 8,491 | (63.2) |
|  | Ex-smoker | 22,879 | (6.6) | 18,524 | (6.5) | 3,148 | (6.7) | 2,409 | (6.8) | 451 | (5.9) | 186 | (6.5) | 102 | (7.6) | 1,207 | (9.0) |
|  | Current smoker | 81,120 | (23.5) | 66,961 | (23.5) | 10,418 | (22.2) | 7,802 | (22.2) | 1,647 | (21.6) | 636 | (22.1) | 333 | (24.7) | 3,741 | (27.8) |
| Smoking status (Men) | Never-smoker | 84,217 | (48.8) | 72,246 | (50.0) | 9,263 | (43.8) | 7,424 | (45.1) | 1,204 | (40.6) | 406 | (36.4) | 229 | (37.1) | 2,708 | (38.3) |
|  | Ex-smoker | 19,610 | (11.4) | 15,679 | (10.9) | 2,849 | (13.5) | 2,192 | (13.3) | 392 | (13.2) | 170 | (15.2) | 95 | (15.4) | 1,082 | (15.3) |
|  | Current smoker | 68,912 | (39.9) | 56,579 | (39.2) | 9,048 | (42.8) | 6,846 | (41.6) | 1,370 | (46.2) | 539 | (48.3) | 293 | (47.5) | 3,285 | (46.4) |
| Smoking status | Never-smoker | 157,657 | (91.1) | 127,671 | (90.6) | 24,203 | (93.5) | 17,559 | (93.7) | 4,306 | (92.8) | 1,654 | (93.6) | 684 | (93.6) | 5,783 | (90.9) |
| (Women) | Ex-smoker | 3,269 | (1.9) | 2,845 | (2.0) | 299 | (1.2) | 217 | (1.2) | 59 | (1.3) | 16 | (0.9) | 7 | (1.0) | 125 | (2.0) |
|  | Current smoker | 12,208 | (7.1) | 10,382 | (7.4) | 1,370 | (5.3) | 956 | (5.1) | 277 | (6.0) | 97 | (5.5) | 40 | (5.5) | 456 | (7.2) |
| Drinking status | Non-drinker | 266,918 | (78.7) | 221,936 | (79.2) | 35,475 | (77.7) | 26,445 | (77.3) | 5,866 | (79.5) | 2,169 | (77.5) | 995 | (76.0) | 9,507 | (71.4) |
|  | Occasional drinker | 39,478 | (11.6) | 32,820 | (11.7) | 4,689 | (10.3) | 3,592 | (10.5) | 696 | (9.4) | 279 | (10.0) | 122 | (9.3) | 1,969 | (14.8) |
|  | Regular drinker | 32,641 | (9.6) | 25,300 | (9.0) | 5,510 | (12.1) | 4,154 | (12.1) | 812 | (11.0) | 352 | (12.6) | 192 | (14.7) | 1,831 | (13.8) |
| Body mass index | <18.5 | 33,308 | (7.2) | 26,577 | (7.4) | 4,406 | (6.7) | 2,853 | (5.9) | 1,009 | (8.9) | 383 | (8.6) | 161 | (8.1) | 2,325 | (6.4) |
|  | 18.5~24 | 293,049 | (63.6) | 234,579 | (65.4) | 35,026 | (53.3) | 25,660 | (53.5) | 5,967 | (52.7) | 2,321 | (52.2) | 1,078 | (54.0) | 23,444 | (64.3) |
|  | 25~29 | 114,257 | (24.8) | 83,464 | (23.3) | 21,552 | (32.8) | 16,101 | (33.6) | 3,471 | (30.7) | 1,367 | (30.8) | 613 | (30.7) | 9,241 | (25.3) |
|  | ≧30 | 20,299 | (4.4) | 14,086 | (3.9) | 4,739 | (7.2) | 3,357 | (7.0) | 865 | (7.6) | 373 | (8.4) | 144 | (7.2) | 1,474 | (4.0) |
| Physical activity | Inactive | 181,794 | (51.1) | 148,488 | (50.9) | 26,108 | (53.1) | 19,155 | (52.1) | 4,444 | (55.5) | 1,707 | (56.1) | 802 | (56.4) | 7,198 | (49.9) |
|  | Low | 85,563 | (24.1) | 72,244 | (24.7) | 10,051 | (20.4) | 7,642 | (20.8) | 1,590 | (19.9) | 553 | (18.2) | 266 | (18.7) | 3,268 | (22.7) |
|  | Moderate | 53,494 | (15.0) | 43,729 | (15.0) | 7,664 | (15.6) | 5,791 | (15.8) | 1,202 | (15.0) | 453 | (14.9) | 218 | (15.3) | 2,101 | (14.6) |
|  | High | 21,816 | (6.1) | 17,013 | (5.8) | 3,565 | (7.2) | 2,716 | (7.4) | 521 | (6.5) | 239 | (7.9) | 89 | (6.3) | 1,238 | (8.6) |
|  | Very high | 12,955 | (3.6) | 10,514 | (3.6) | 1,819 | (3.7) | 1,430 | (3.9) | 250 | (3.1) | 91 | (3.0) | 48 | (3.4) | 622 | (4.3) |
| Hypertension | None | 367,272 | (79.7) | 298,479 | (83.2) | 42,021 | (63.9) | 30,372 | (63.3) | 7,617 | (67.2) | 2,838 | (63.8) | 1,194 | (59.6) | 26,772 | (73.4) |
|  | Yes | 93,759 | (20.3) | 60,276 | (16.8) | 23,760 | (36.1) | 17,625 | (36.7) | 3,711 | (32.8) | 1,613 | (36.2) | 811 | (40.4) | 9,723 | (26.6) |
| Diabetes | None | 435,832 | (94.5) | 343,469 | (95.7) | 58,235 | (88.5) | 42,390 | (88.3) | 10,148 | (89.6) | 3,949 | (88.7) | 1,748 | (87.2) | 34,128 | (93.5) |
|  | Yes | 25,199 | (5.5) | 15,286 | (4.3) | 7,546 | (11.5) | 5,607 | (11.7) | 1,180 | (10.4) | 502 | (11.3) | 257 | (12.8) | 2,367 | (6.5) |
| Cough recently | None | 413,893 | (89.8) | 323,143 | (90.1) | 57,514 | (87.4) | 42,195 | (87.9) | 9,865 | (87.1) | 3,817 | (85.8) | 1,637 | (81.6) | 33,236 | (91.1) |
|  | Yes | 47,138 | (10.2) | 35,612 | (9.9) | 8,267 | (12.6) | 5,802 | (12.1) | 1,463 | (12.9) | 634 | (14.2) | 368 | (18.4) | 3,259 | (8.9) |
| Blood in sputum | None | 450,356 | (97.7) | 349,962 | (97.5) | 64,331 | (97.8) | 46,990 | (97.9) | 11,055 | (97.6) | 4,342 | (97.6) | 1,944 | (97.0) | 36,063 | (98.8) |
|  | Yes | 10,675 | (2.3) | 8,793 | (2.5) | 1,450 | (2.2) | 1,007 | (2.1) | 273 | (2.4) | 109 | (2.4) | 61 | (3.0) | 432 | (1.2) |
| Wheezing | None | 427,607 | (92.8) | 334,476 | (93.2) | 59,050 | (89.8) | 43,625 | (90.9) | 10,044 | (88.7) | 3,783 | (85.0) | 1,598 | (79.7) | 34,081 | (93.4) |
|  | Yes | 33,424 | (7.2) | 24,279 | (6.8) | 6,731 | (10.2) | 4,372 | (9.1) | 1,284 | (11.3) | 668 | (15.0) | 407 | (20.3) | 2,414 | (6.6) |
| Asthma history or medication | None | 448,815 | (97.4) | 350,276 | (97.6) | 63,520 | (96.6) | 46,815 | (97.5) | 10,813 | (95.5) | 4,141 | (93.0) | 1,751 | (87.3) | 35,019 | (96.0) |
|  | Yes | 12,216 | (2.6) | 8,479 | (2.4) | 2,261 | (3.4) | 1,182 | (2.5) | 515 | (4.5) | 310 | (7.0) | 254 | (12.7) | 1,476 | (4.0) |
| * N Number of participants  PRISm: Preserved Ratio Impaired Spirometry; COPD: Chronic Obstructive Pulmonary Disease. | | | | | | | | | | | | | | | | | |
