## Supplementary figures and images for "Impaired lung function and lung cancer risk in 461,183 healthy individuals: a cohort study"

### Supplementary Figure 1

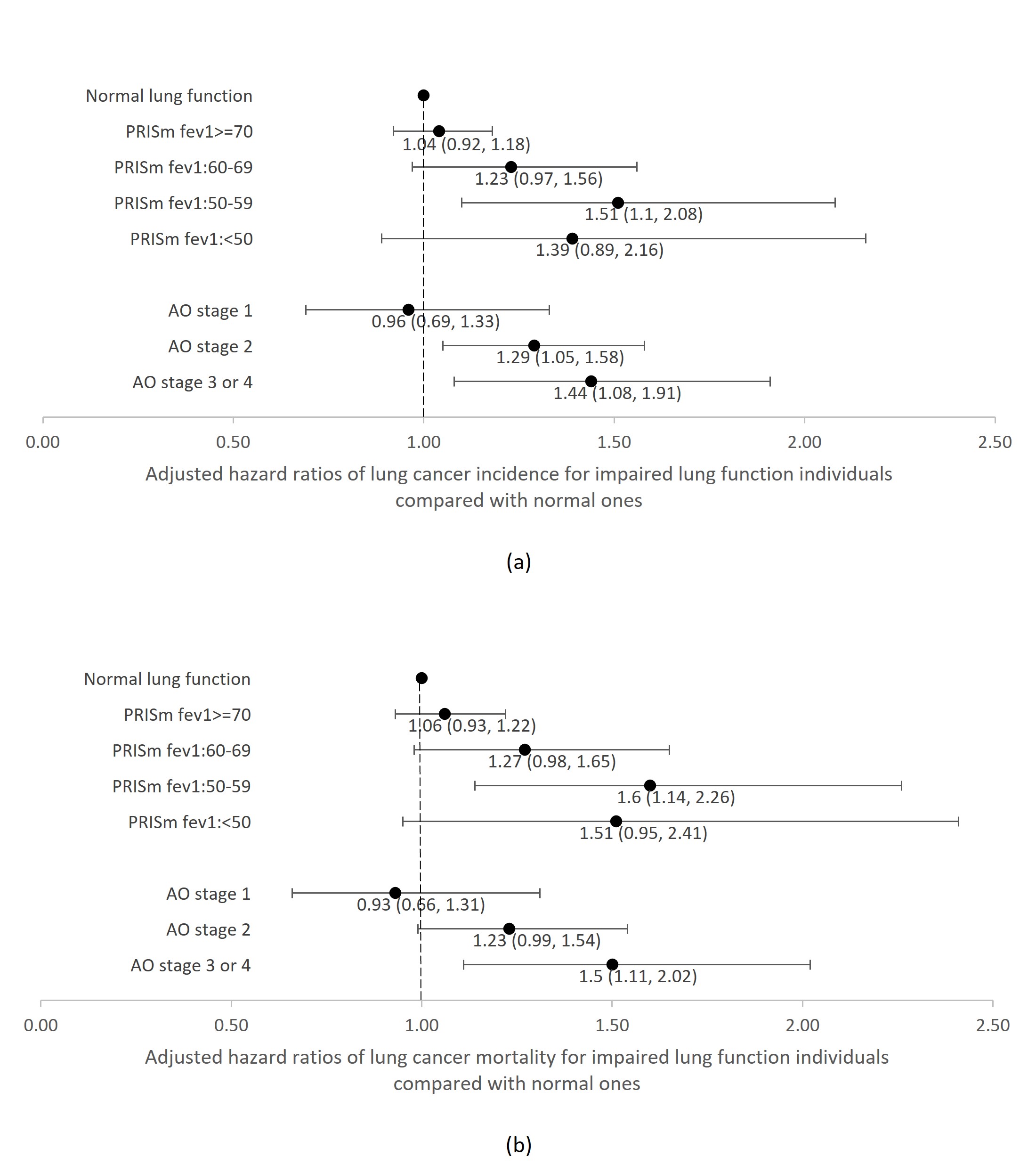

### Supplementary Figure 2

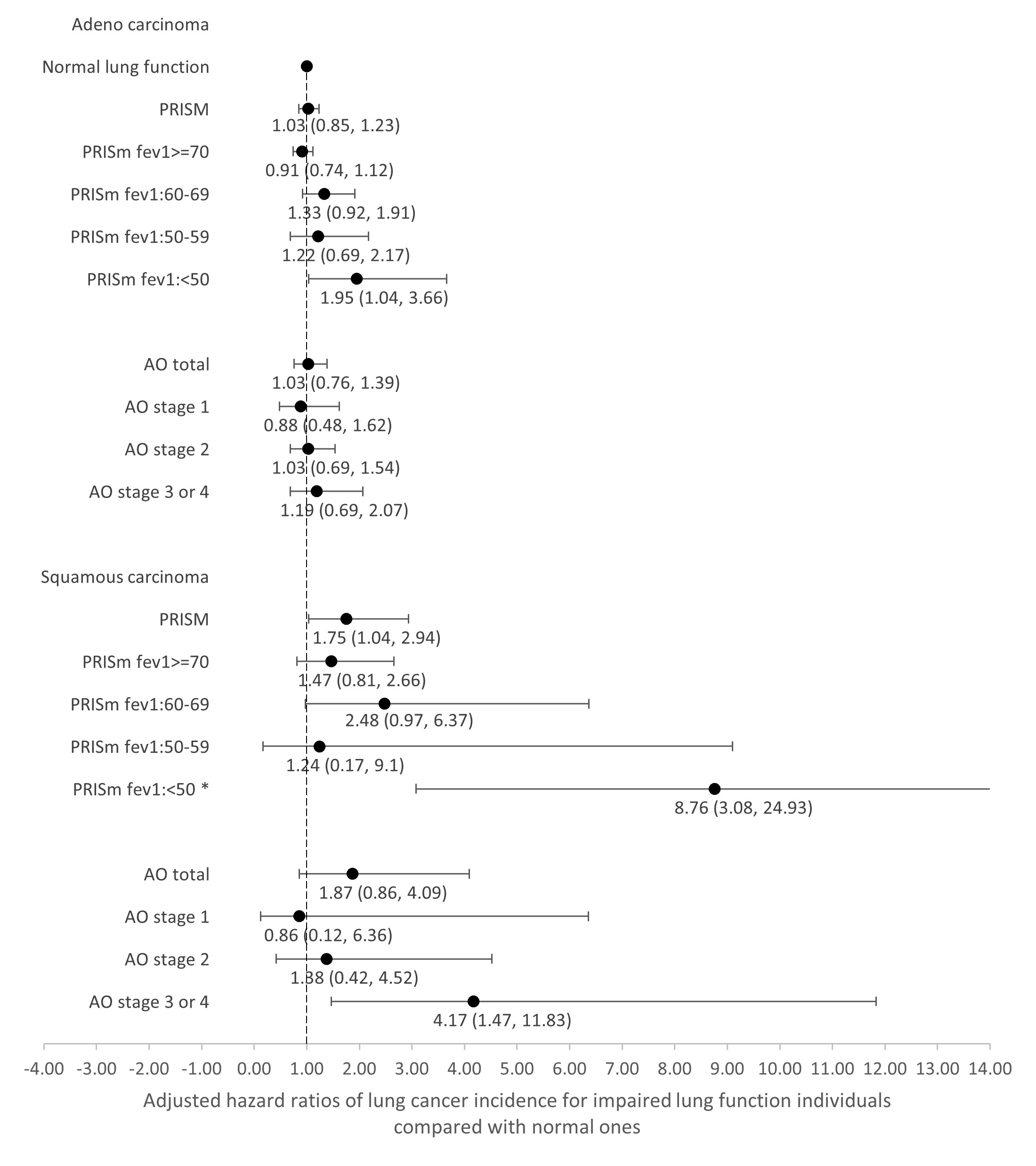
